# Exploration of post-insertion challenges edentulous patients present to dental practitioners during removable complete denture rehabilitation in Makerere University Dental Hospital in Uganda

**DOI:** 10.1101/2024.12.21.24319253

**Authors:** David Nono, Godfrey Bagenda, Isaac Okullo, Charles Mugisha Rwenyonyi

## Abstract

**Background:** Edentulism in a major global burden that contributes to disability and impairment. Globally, edentulous patients frequently receive removable complete dentures (RCDs) and though crucial, effective adaptation is still difficult. RCD remains a preferred treatment modality for edentulous patients worldwide. The frequency of full edentulism varies both within and between countries. In Uganda, 1.8% of people aged 20 years and above are edentulous. Despite advances in dental technology and material science, the successful adaptation of RCDs remains a challenge for both patients and dental professionals. Our present study aimed to explore the post-insertion challenges patients presented to dental practitioners during their rehabilitation with removable complete dentures in Makerere University Dental Hospital.

**Methods:** This was a qualitative study using a phenomenological technique and purposive sampling to select 25 participants. After obtaining institutional review board approval and written informed consent, semi-structured in-depth and key informant interviews of dental practitioners were conducted and the data were analyzed thematically.

**Results:** It was revealed that patients with RCDs commonly reported substantial post-insertion challenges like pain and discomfort, drooling, difficulty speaking, eating, oral hygiene and salivating. Patients’ confidence and self-esteem were severely influenced by their displeasure with the way their dentures looked. Furthermore, patients frequently had denture instability. In addition, patients received helpful post-insertion advice that emphasized the significance of oral health education and support from dental practitioners. This advice included instructions on denture cleaning, maintenance and adaption.

**Conclusion:** The present study identified several post-insertion challenges faced by patients after receiving RCDs. It is important to provide integrated support for patients and practitioners to enhance effective RCD therapy.

## Background

Tooth loss (edentulism) continues to be a major health burden globally, particularly for the elderly. Although the occurrence of complete edentulism has declined in recent years, it is still a crippling and permanent disorder. It is frequently referred to as the “final marker of disease burden for oral health”[1]. The frequency of total tooth loss varies both within and between countries. In Uganda, 1.8% of people aged 20 years and above have lost teeth [2]. Total tooth loss leads to handicap, incapacity and disability [3].

Globally, removable complete dentures (RCDs) remain a preferred treatment modality for edentulous patients [4]. Despite advances in dental technology and material science, the successful adaptation of RCDs remains a challenge for both patients and dental professionals. While much attention has been given to the fabrication and initial fitting of RCDs, the post-insertion phase is equally critical for ensuring patient satisfaction and long-term success [5]. The most challenging aspect of dental treatment in day-to-day practice for most dental practitioners remains the successful management of complete denture patients who often encounter recurrent difficulties with their prostheses. The critical factor being patient adaptation to RPD. However, while many patients with a positive perspective overcome hurdles during and after treatment, some are unable to adapt physically or psychologically, particularly with advancing age. Often, this is due to unrealistic expectations resulting in denture post-insertion complaints such as discomfort, pain, difficulty in mastication, slurred speech and aesthetic concerns. This highlights the importance of early issue identification and fostering a positive patient-prosthodontist relationship [5–8].

Despite the importance of post-insertion care in ensuring edentulous patient satisfaction and treatment success, there is no published literature on post-insertion challenges faced by edentulous patients in Uganda. The study aimed to explore post-insertion challenges edentulous patients presented to dental practitioners during removable complete denture rehabilitation in Makerere University Dental Hospital in Uganda.

## Materials and methods

### Study design

The study employed a qualitative method of data collection using in-depth and key informant interviews of dental practitioners.

### Study site

The study was conducted in Makerere University Dental Hospital in Kampala. Kampala is the capital city of Uganda. The hospital is a teaching and healthcare service delivery facility of Makerere University. It is the largest and adequately equipped dental facility employing the highest number of dental specialists in Uganda. It has a well-established prosthetic dental laboratory and provides specialized dental services such as the rehabilitation of edentulous patients with RCD who comprise staff and students of the university as well as the surrounding community. Every month, the hospital attends to about 660 outpatients of whom 20 are rehabilitated with RCD (Registry of Dental Records, 2022).

### Selection of Study Participants

Key informants were purposively selected based on their expertise in the clinical services. To guarantee an equitable representation of the study population, the selection process also took into account differences in the length of practice level of training, roles in denture manufacturing processes and fitting. Based on data saturation, 12 technologists and 13 dental surgeons were interviewed as key informants.

### Inclusion criteria

Dental surgeons and dental technologists participating in the provision of removable complete dentures in Makerere University Hospital who were willing to participate in the study.

### Exclusion criteria

Dental technologists and dental surgeons who were ill or absent were excluded from the study.

### Data Collection

The participants gave written informed consent before taking part in the study. They were assured of confidentiality where names or other identifying information were not used when gathering data or writing reports. The interviews took place in a private setting favorable to the participants to freely share their perspectives. A semi-structured interview was conducted by the principal investigator (DN) using a guide while the research assistant took notes and audio recorded the interview. The interview, which lasted between 30 and 40 minutes covered themes related to various elements of post-insertion challenges patients presented to dental practitioners during RCD therapy. The process of gathering and analyzing data was participatory. Informational redundancy or the absence of new information from the interview determined the final participant [9–10]. The recruitment of participants started on 15^th^ September 2023 and ended on 20^th^ November 2023.

### Data Management and Analysis

Audio recording was transcribed verbatim to form a code book. Five transcripts were used for testing the code book, which was then uploaded into Nvivo 14 for methodical organization and analysis. Themes were used to analyze the data [11]. Emerging and recurring themes were identified and analyzed after reading and re-reading the transcripts. Individual quotes were used to represent challenges of patients during post-insertion of RCDs.

### Quality control

The principal investigator (DN) pretested the data collection instruments and made modifications to increase their validity and reliability. The research assistant was trained by DN in data collection in addition to his background in the social sciences and expertise in qualitative research methods. Interviews were audio-recorded to capture any conversation that might have been missed during note-taking. Extra notes that captured gestures and body language during the interviews were also made. Four criteria were used to ensure reliability: transferability, credibility, confirmability and dependability. Regarding transferability, a detailed description of the research context, the characteristics of the selected participants and settings, and how they influenced the findings was provided. Credibility was established through peer debriefing and the involvement of more seasoned qualitative researchers who assessed and offered input on the study design and results to ensure that the data was accurate and relevant. For dependability, an elaborate description of the steps and methods used in data collection and analysis have been indicated to allow replication of the findings. To allow for confirmability, a description of the data analysis and presentation of findings has been provided. A well-defined coding scheme was utilized to generate codes and discover trends in the analysis, ensuring that the conclusions of the study were objective and based on the testimonies and statements of the participants.

### Ethical Considerations

The study protocol was approved by Makerere University School of Health Sciences Research Ethics Committee (Reference Number: MAKSHSREC-2023-486) and Uganda National Council for Science and Technology (Reference Number: HS3092ES). The administration of the Makerere University Dental Hospital granted permission to conduct the study. Written informed consent was obtained from all respondents who participated in the study. The participants were informed of the study’s objectives and were asked to voluntarily participate in accordance with the Helsinki Declaration [12].

## Results

### Socio-demographic characteristics

Twenty five dental practitioners (12 dental technologists and 13 dental surgeons) participated in the study and their demographic characteristics varied (Table 1). The participants, respectively, held Bachelor of Dental Technology and Bachelor of Dental Surgery. Eighteen participants were males and more than half (n=14) were aged between 26 and 35 years (Table 1).

**Table 1.**
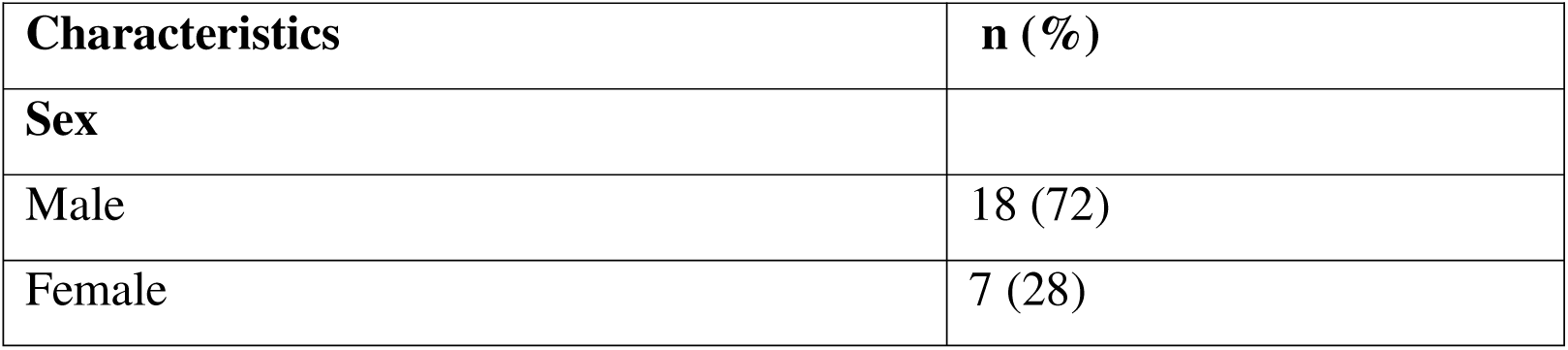
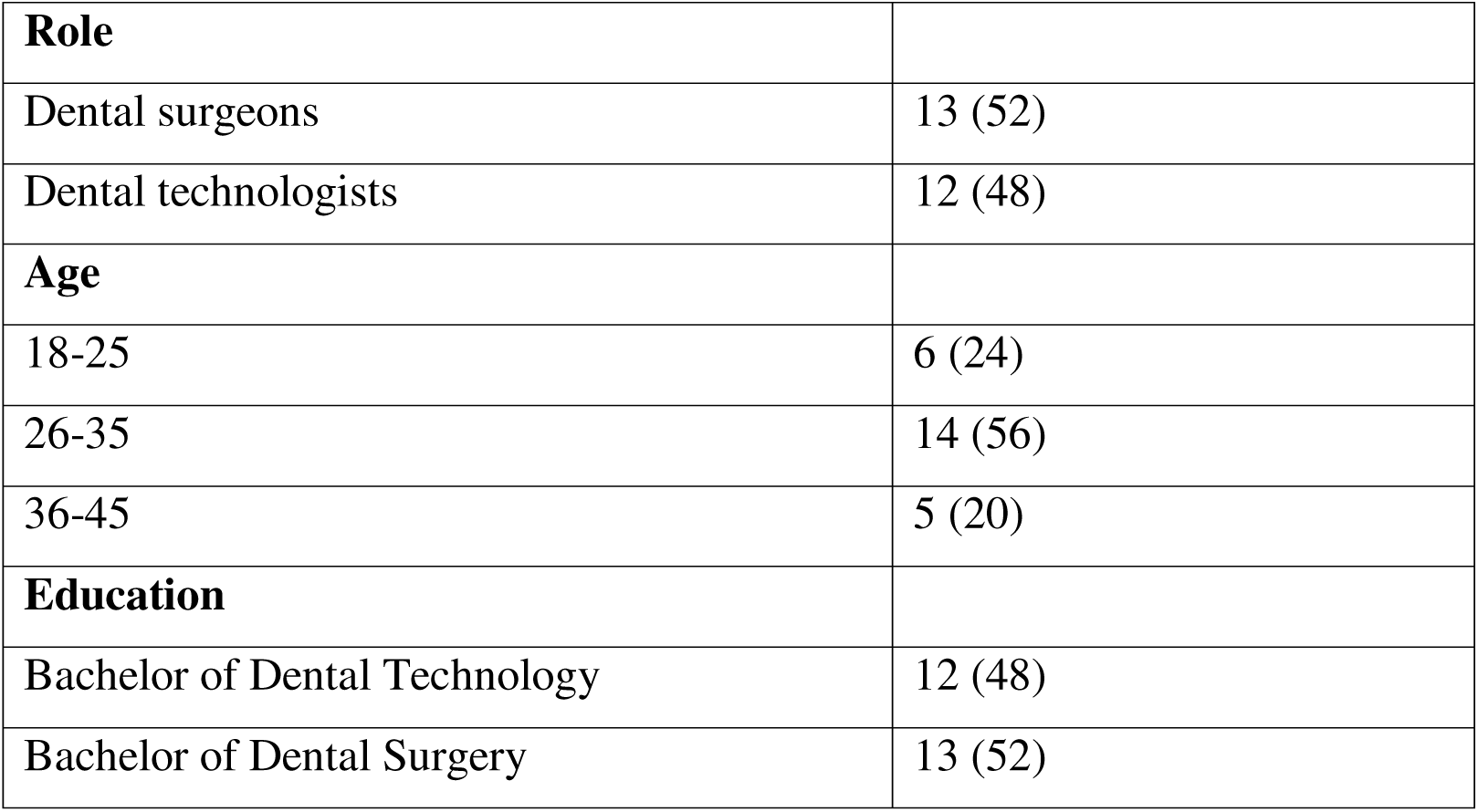
The frequency distribution of the respondents according to their social demographic characteristics (N=25)

**Table 2:** Themes and subthemes.

| **Themes** | **Subthemes** |
| --- | --- |
| Challenges faced by patients after delivery of removable complete denture (RCD) | • Difficulty in using the RCD<br>• Deteriorating hygiene, ulceration and pain<br>• Patients' unrealistic expectations<br>• Removable complete dentures are not stable |
| Removable complete denture post-insertion advice provided | • Hygiene and routine cleaning care for RCD<br>• Speech impairment, pain and discomfort |

### Challenges faced by patients after delivery of removable complete denture (RCD)

We asked dental practitioners about the challenges that patients faced as a result of wearing RCDs. Their narratives revealed a number of challenges related to difficulty in speech, eating and saliva drooling out of mouths, dentures being expensive, hygiene, ulceration and pain while some revealed complete dentures not being stable and falling out.

#### Difficulty in using the RCD

Participants reported that their clients expressed difficulty in speech, eating and some had saliva drooling out of mouths as a result of using dentures. As part of challenges in adaptation, some participants revealed how difficult it becomes for patients to chew food normally because in the first days, the gum got irritated by the sharp edges of the RCDs if not well polished/finished. Others lose a sense of taste in the mouth.

> “They reported that their gums were irritated by the dentures’ sharp edges. There are those who said they cannot use RCDs for eating” (P003_Male).

> “Many elderly patients came back and said ‘Doctor, I no longer have a taste for my food’. Then, I just counselled them that the denture tends to cover the taste buds. So, it hinders their taste” (P017_33 Male).

Some patients also experienced saliva drooling out of the mouth especially in the first days after RCD delivery is done and as a result, speaking becomes difficult. And the drooling out of too much saliva is associated with inexperience to control the tongue in the mouth.

> “Speaking was very difficult in the first few weeks. So, basically saliva was drooling out of the mouth and all that” (P004_Male).

> “The tongue was usually less controlled and they have a tendency of allowing saliva to spill out” (P002_Female).

RCDs were also associated with speech impairment. This was a situation where some patients failed to pronounce certain words correctly as they used to before.

> “Maybe some little bit of speech impairment. So, for them they felt like they were not talking so well and having problems of pronouncing words like S, T, and H” (P003_Male).

#### Deteriorating hygiene, ulceration and pain

The pain due to the wounds caused by the sharp edges of the poorly polished removable complete dentures were also reported as challenges faced by the patients. After a few days of complete denture procedures, dental practitioners reported to have received patients who come back complaining of ulcers in the mouth.

> “In the first one or two weeks, there are ulcers that come with wearing removable complete dentures and those ulcers are basically because of high occlusion contacts of complete dentures and a client was not comfortable when an ulcer was in the mouth. It is very hard for a client to be comfortable, so those wounds come as a side effect in the mouth…” (P004_Male).

The dental practitioners thought that the ulcers were caused by extensions that were not well polished to the extent that they scratch the soft tissues in the mouth, hence a patient developed wounds and eventual pain.

> “They can occasionally arrive with ulcerations from extensions if they were not properly polished or if anything sharp had scratched the soft tissues within the mouth.” (P013_Female).

Similarly, poor hygiene was mentioned as one of the challenges patients face. One participant explained this in detail:

> “Some patients could easily get candida depending on the oral hygiene. Some got issues with chewing, saliva drooling out of the mouth and speech problems in the first few weeks” (P004_Male).

#### Patients’ unrealistic expectations

As a result of having the fabricated RCDs different from their lost teeth, patients feel disappointed and don’t feel like wearing the dentures again to ensure that other people don’t get to know that one is wearing a RCD.

> “I delivered a removable complete denture to the patient, but came back and said that he didn’t want those teeth [denture] anymore because people have realized they are just artificial teeth.” (P001_Male).

> “Most times patients thought that they are going to be given like permanent fixed complete dentures which was not the case” (P013_Female).

#### Removable complete dentures are not stable

Some participants reported that patients faced a challenge of dentures not being stable and sometimes keep shifting and even falling out, which was discomforting and took time for patients to get used to. However, with time the patients got used and learnt how to control their muscles so that the dentures were held firm on the jaws/ridge.

> “There were times when the complete dentures were unstable and moved sideways when chewing hence unable to bite” (P002_Female).

> “There was a patient whose denture was falling even after adjusting the flanges, but eventually it stabilized. I think it was because they did not know how to control muscles to retain them [dentures]. So, they were still adapting to them” (P015_Male).

### Removable complete denture post –insertion advice provided

We took interest in knowing if there was any advice or support given to the patients after the delivery of the removable complete dentures. Our findings revealed that patients had received a range of post-insertion advice from the dental practitioners.

#### Hygiene and routine cleaning care for RCD

The common advice provided to patients included routine care by brushing every morning and evening as well as advice on the procedures and precautions during cleaning. In some cases, the advice was not to use a tooth brush and paste, but to brush with a clean cloth with soap and clean water depending on the reviews done. Another participant emphasized the care received by revealing that they advised the patients to take extra care of their complete dentures just as they take care of the rest of their body parts like the eyes.

> “The routine is brushing after every meal in the morning and in the evening. You can get a clean cloth with soap and clean water and try to clean inside and outside, and don’t allow any food to get stuck leading plague accumulating that will affect the retention of the complete denture.” (P001_Male).

> “At night when they are going to sleep, they should clean the dentures with a brush and soap, and put them in a container with clean cold water. In the morning, they should wear them and eat breakfast and thereafter remove them from the mouth, brush their mouth and clean the complete dentures.” (P007_Male).

The intention of all the forms of post-insertion advice given, especially the cleaning and caring of the complete dentures is to prevent injuries to the soft tissues that are likely to lead to infection due to microorganisms accumulating on food particles on the uncleaned complete dentures. Some of the dental practitioners narrated that oral hygiene was very important to prevent oral infections:

> “I tell them that these removable complete dentures sometimes get the microorganisms attached to them. So, if they over build, they might cause soft tissue infections in oral cavity. So, after some days of wearing denture, if they drop them (dentures) into liquid soap solution overnight, my assumption is that the microorganisms will die” (P008_Male).

The patients were also advised not to wear the dentures at night because the RCDs tend to create pressure on the gum and eventually develop painful sores.

> “..…so we encourage them (patients) to remove them (dentures) from the mouth at night and keep them in cold water in order to relieve pressure on the gum” (P003_Male).

#### Speech impairment, pain and discomfort

The other advice given to patients was related to the kind of impact of the complete dentures on the patient’s normal routine life, for example, the advice that the dentures may cause some speech impairments, pain and discomfort, which are temporally and they will eventually get used to and normalize.

> “For speech related issues, I tell them that it is normal to experience speech impairment, but with time, they will get used to them and speak normally” (P013_Femal).

They also informed the patients about the inconveniences that may come just after the complete denture delivery, which may discourage them to keep wearing them, but instead remove and stop putting them on. They encouraged the patients to keep wearing their dentures despite all inconveniences and assured them that they will eventually get used and become comfortable and confident.

> “Put it on, don’t keep out, you may be uncomfortable on day one or two, but maintain it on and later, you will gain confidence and be comfortable with it” (P011_Female).

## Discussion

The present qualitative study used a phenomenological approach^13^ to look into the post-insertion challenges that patients presented to dental practitioners during the rehabilitation using RCDs. In addition, guiding policymakers to evaluate the current recommendations for fabricating RCDs [14] that are appropriate for specific edentulous patients in Uganda, the findings of the present study could be utilized in training of dental students as well as catalyst for further studies. However, the objective was to investigate the post-insertion difficulties that patients report to dental practitioners during their rehabilitation with removable complete dentures at the Makerere University Dental Hospital in Uganda. We are not aware of any published information about the difficulties Ugandan edentulous patients encounter following insertion of RPDs. The current study contributed to the growing literature about the post-insertion difficulties that edentulous patients shared with the dental practitioners in elsewhere in the world.

The present study revealed that patients present with various post-insertion challenges after receiving RCDs. The challenges included difficulties in speech, eating and saliva drooling, which is consistent with findings from a previous survey [15]. However, Gosavi et al. [15] also highlighted additional issues such as bad breath, tongue restriction and food accumulation, which were not explicitly reported in the present study. Additionally, the present study indicated that poorly polished RCDs can lead to discomfort and pain for patients, which is consistent with findings in a previous study [16].

Another significant challenge reported in the present study was poor oral hygiene among patients using RCDs. A study conducted by Dakka et al. [17] found that poor oral hygiene in the elderly population using removable dentures led to serious complications such as biofilm formation, pneumonia, stomatitis and accidental ingestion/aspiration of dentures, indicating the need for better education, guidance and access to dental care to improve oral hygiene and overall well-being of the denture wearing population [17]. Patients often felt disappointed when the fabricated denture teeth did not match their lost natural teeth, leading to reluctance in maintaining proper oral hygiene and a general resistance to the lifestyle changes associated with wearing dentures. This dissatisfaction with the appearance of their dentures could have a significant impact on patients’ confidence and self-esteem, leading to social and psychological consequences as earlier mentioned [18].

In the present study, some participants reported denture instability as a common challenge faced by patients wearing RCDs. Dentures reportedly shifted or even fell out, causing discomfort and requiring time to adjust. However, over time, patients typically learnt how to control their muscles to stabilize the dentures in their oral cavities. This finding corroborates a previous study [19]. Some participants reported that patients complained of the prohibitively high cost of paying for RCDs. Additionally, the alternative option of using implants for denture stability was even more expensive for the patients, similar to a report from Russo et al. [20]

In the present study, some participants advised patients to use a clean cloth with soap and water and not necessarily a toothbrush and toothpaste to clean their dentures, but in any case, depending on individual circumstances. This recommendation partly corroborates other findings which have shown that mechanical cleaning with a soft toothbrush and mild soap is effective in removing plaque and debris from denture surfaces [4, 21]. Participants emphasized the importance of advising patients to take extra care in looking after their complete dentures, similar to caring for other body parts. Patients were also provided with guidance on how to store their dentures, particularly at night, to prevent them from warping.

The present study revealed that patients were informed about the potential impact of complete dentures on their normal routine life. They were advised that dentures may initially cause speech impairment, pain and discomfort, which are usually temporary as they will eventually adapt and normalize. Participants encouraged patients to persevere and continue wearing their dentures, assuring them that they would eventually become comfortable and confident. This was in line with other authors [5–8] who highlighted how many patients with a positive perspectives overcame post-insertion hurdles like discomfort, pain, difficulty in mastication, slurred speech and aesthetic concerns following RCD treatment. However, those with unrealistic expectations ended up with prolonged post-insertion denture complaints such as discomfort, pain, difficulty in mastication, slurred speech and aesthetic.

### Recommendations

- Dental practitioners should focus on ensuring proper fitting and polishing of RCDs to minimize discomfort and pain for patients. This includes attention to detail during the fabrication process to avoid sharp edges and irregularities that can cause irritation to the oral tissues.
- There is need for better education and guidance on oral hygiene practices among patients wearing RCDs. Dental practitioners should provide comprehensive instructions on denture cleaning and maintenance to prevent subsequent infections.
- Dental practitioners should provide thorough counseling and support to patients undergoing RCD treatment to address patients’ expectations and concerns through reassurance during the adaptation period.

### Implications for Clinical Practice

- Dental practitioners should pay close attention to the fitting and polishing of RCDs to minimize discomfort and pain for patients.
- There is a need for better education and guidance on oral hygiene practices among patients wearing RCDs.
- Dental practitioners should provide thorough counseling and support to patients undergoing RCD treatment.

### Implications for Future Research

- Future research should focus on investigating RPD post insertion challenges from patients themselves as opposed to secondary information from dental practitioners.
- There is a need to explore the impact of RCDs on patients’ overall well-being and quality of life.

### Study Limitations

- The study research relied on subjective/secondary information from dental practitioners, which may have been influenced by recall bias and perspectives. However, efforts were made to minimize this bias through rigorous data analysis and triangulation of findings.
- This was a qualitative study carried out in one dental hospital, which needs caution when generalizing to a Ugandan population.

## Conclusion

Our study identified several post-insertion challenges faced by patients after receiving RCDs, including difficulties in speech, eating and saliva drooling as well as poor oral hygiene. It is important to provide integrated support to patients and dental practitioners to enhance effective RCD therapy. This would go a long way to improve the stability and affordability of RCDs and provide guidance on denture cleaning, maintenance and adaptation.

## Declaration

### Ethics approval and consent to participate

Both Makerere University School of Health Sciences Research and Ethics Committee (Reference Number: MAKSHSREC-2023-486) and Uganda National Council for Science and Technology (Reference Number: HS3092ES) provided ethical approval of the protocol. The Makerere University Dental Hospital administration granted permission to conduct the study. In compliance with the Helsinki Declaration, all respondents provided written informed consent prior to their participation in the study [12]. Only the investigators had access to the cabinet containing all of the collected data kept under lock and key.

### Consent for publication

Not applicable

### Availability of data and materials

All relevant data are within the manuscript and its supporting information file.

## Supporting information

S1 File: Interview guide for dental surgeons and dental technologists

## Data Availability

All relevant data are within the manuscript and its supporting information file.

## Acknowledgments

The authors are grateful to the Government of Uganda through the MAK-RIF secretariat for financially supporting this study and participants for their willingness to participate in the study.

## Author’s contributions

**Conceptualization of study:** David Nono, Godfrey Bagenda, Isaac Okullo, Charles Mugisha Rwenyonyi

**Data curation:** David Nono and Godfrey Bagenda

**Formal analysis:** David Nono, Godfrey Bagenda, Isaac Okullo, Charles Mugisha Rwenyonyi

**Funding acquisition:** David Nono, Godfrey Bagenda, Charles Mugisha Rwenyonyi

**Investigation:** David Nono, Charles Mugisha Rwenyonyi

**Methodology:** David Nono, Godfrey Bagenda, Isaac Okullo, Charles Mugisha Rwenyonyi

**Project administration:** David Nono

**Software:** David Nono, Godfrey Bagenda

**Supervision:** Godfrey Bagenda, Isaac Okullo, Charles Mugisha Rwenyonyi.

**Validation:** Charles Mugisha Rwenyonyi

**Visualization:** David Nono, Godfrey Bagenda, Charles Mugisha Rwenyonyi

**Writing original manuscript draft:** David Nono

**Reviewing & editing the manuscript:** David Nono, Godfrey Bagenda, Isaac Okullo, Charles Mugisha Rwenyonyi

